# Learning to Treat Hypotensive Episodes in Sepsis Patients Using a Counterfactual Reasoning Framework

**DOI:** 10.1101/2021.03.03.21252863

**Authors:** Russell Jeter, Li-Wei Lehman, Christopher Josef, Supreeth Shashikumar, Shamim Nemati

## Abstract

The optimal treatment strategy for volume resuscitation and vasopressor dosing to combat hypotensive episodes in septic patients remains a subject of ongoing controversy and can vary from clinician to clinician. We develop a machine learning approach to guide a fluid and vasopressor dosing strategy that adapts to patient-specific clinical states to improve the survival of septic patients. We adopt a model-free reinforcement learning (RL) framework in a continuous action space with a clinically significant reward function, and use a Switching Generalized Linear Model (SGLM) to characterize patient-specific clinical states. We use retrospective data from the MIMIC III database to train this model to learn volume resuscitation and vasopressor dosing strategies among the 5,366 patients (totalling 352,328 unique hourly measurements) with ICU-onset sepsis or septic shock, as diagnosed by the Sepsis-3 definition. The RL agent receives short- and long-term rewards associated with optimizing in-hospital survival and avoiding end-organ damage to learn volume resuscitation and vasopressor dosing strategies. On average, the RL agent learns to resuscitate patients earlier than clinicians with a fluid bolus (one hour vs. four hours after the diagnosis of sepsis), and improves the expected survival by ≈ 3%. Our preliminary results indicate that adherence to RL-based individualized fluid and vasopressor dosing recommendations is associated with a significant mortality reduction in septic patients, even after adjusting for severity of illness.

## I. Introduction

Sepsis is a life-threatening condition that arises when the body’s response to infection injures its own tissues [1]. Sepsis is diagnosed when there is a suspected infection (indicated by the administration of antibiotics or collection of cultures) and a two-point increase in the Sequential Organ Failure Assessment Score (SOFA-Score) [1]. In the US nearly 6% of all hospitalized patients carry a diagnosis of sepsis. When all hospital deaths are considered nearly 35% can be attributed to sepsis [2].

Even when sepsis is recognized in a timely manner it can proceed to septic shock. Shankar-Hari et al. define septic shock as a subset of sepsis in which underlying circulatory, cellular, and metabolic abnormalities are associated with a greater risk of mortality than sepsis alone [3]. The Sepsis-3 Guidelines for diagnosis of septic shock are hypotension (abnormally low blood pressure) requiring therapeutic intervention to maintain mean arterial blood pressure (MAP) of 65 mmHg or greater [1] and lactate > 2mmol/L. While there is a general consensus regarding the first hour of treatment, there exists no dosing or prescribing guidelines beyond the recommendation to maintain a MAP ≥ 65 mmHg. The use of vasopressors and IV fluids can vary according to concomitant patient comorbidities, a clinician’s area of expertise, and institutional policies making it difficult to ascertain which combination of vasopressors and IV fluids is ideal for septic patients during hypotensive episodes. As such, the optimal treatment strategy for volume resuscitation and vasopressor dosing to enhance outcomes of sepsis remains the subject of ongoing controversy.

Medication dosing in a critical care setting is a complex sequential decision making process that depends not only on a patient’s clinical course, but also unique multifactorial covariates. Moreover, due to the intermittent nature of laboratory measurements clinicians have to deal with delayed feedback and the problem of credit assignment (i.e., deciding which of the many treatments will stabilize the patient). Reinforcement learning provides a natural solution to these types of sequential decision making problems [4]. Based on the patient state (measured by vital signs and clinical tests) and covariate factors (age, gender, etc.) an agent can provide treatment recommendations to clinicians (i.e., what amount of intraveneous fluids to provide to a septic patient to prevent their blood pressure from dropping). Indeed, reinforcement learning has begun to appear in the clinical machine learning literature. Examples include balancing anesthesia levels [5], blood thinner dosing [6], training an artificial pancreas [7], and optimizing mechanical ventilators [8]. Recently, a set of guidelines [9] for the treatment of machine learning in medicine appeared in *Nature Medicine*.

Formally, a reinforcement learning setup consists of an agent placed in some Markov Decision Process (MDP) environment composed of a set of states, 𝒮= {*s*_0_, *s*_1_, …}, a set of actions for the agent, 𝒜 = {*a*_0_, *a*_1_, …}, transition probabilities *P* (*s*^*′*^ |*s, a*) (that is, the probability of the next state *s*^*′*^ given the current state *s* and action *a*), and a reward function *r*(*s, a*) that depends on a given state-action pair. In the setting of dosing septic patients with vasopressors and fluids, for patient *k* the state at each time point *t* is represented by 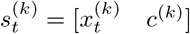, where 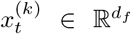 is a vector of the observations (vital signs and clinical variables) and 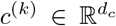 is a vector of the covariates. For each state *s*_*i*_, the agent takes an action *a* ∈ ℝ^2^, the dose pair (*a*^fluid^, *a*^pressor^) of intravenous fluids and vasopressors. By tracking the reward function *r*(*s, a*), the agent learns a *policy* denoted by *π*, which associates a given action *a* with each state *s* that will yield the best (in the experience of the agent) reward.

Traditionally, the components of a reinforcement learning environment are fairly obvious. Consider for example the now-classic RL setting of training an agent to play Atari games [10]. The state consists of the pixels that compose the game screen for a given frame, the actions are the possible game controller options (up, down, left, right, and pressing the action button), and the reward is the change in the score corresponding to a given state and action pair.

The critical care setting provides multiple challenges to the reinforcement learning framework. The first issue is that with few exceptions, there is no *in silico* test-bed in which an agent can freely explore the state-action space by testing various actions on simulated patients. In other words, an agent can only use real, retrospective action data to form its action policy, it can not test actions for itself and see what consequences come from those actions. The most fundamental issue pertains to quantifying a “reward” from the clinical setting. In general, the most natural “reward” in this setting is the patient becoming healthier in response to a treatment, however this is difficult to objectively quantify and usually occurs on a larger time-scale than the observational data is presented.

The layout of this paper is as follows. In Section 2, first we briefly review the critical care dataset used in this study. Then, we detail the methods used for training the RL agent and the state space model. After that, we outline the methods we use to evaluate the RL agent’s policy using counterfactual reasoning and causal treatment effects analysis. In Section 3, we review the results obtained by our model and discuss how our full model compares to less complicated versions of our model (such as a model that learns only from short-term rewards, or a model that does not estimate the state space as well). Lastly, in Section 4, we discuss how our work compares to similar work in the literature, and the implications of our work to the treatment of septic patients. Finally, we highlight meaningful extensions to this work.

## II. Materials and Methods

### A. Dataset

We use retrospective data from the MIMIC III database [11] to develop a model-free reinforcement learning approach to volume resuscitation. Clinical and physiological variables (e.g., laboratory values and vital signs — see the Appendix for a list of the variables included) from the onset of sepsis and the proceeding 72-hour observation window (if available) was utilized in a Switching Generalized Linear Model (SGLM) framework to classify patient-specific clinical states. Our RL agent receives short- and long-term rewards associated with avoiding hypotensive episodes and optimizing in-hospital survival, respectively, among 5,366 patients with ICU-onset sepsis (as defined by the Sepsis-3 criteria).

Clinicians recognize that blood pressure depends on three factors: cardiac output (CO), the diameter of the arteries, and the quantity of fluid inside the vasculature. In critically ill hypotensive patients, vasopressors are routinely used to increase CO and decrease arterial diameter. Additionally, the administration of intravenous (IV) fluids positively influences MAP by increasing the total circulating volume. We utilize fluid administration and normalized vasopressor dosages in each hour to define a continuous two-dimensional set of actions. Age, gender, Elixhauser comorbidity index, and Charlson Comorbidity Index were included as covariates to adjust for severity of illness of each patient. Additional discussion of the dataset is included in the Appendix.

### B. RL Dosing Agent

We seek to construct a RL agent that utilizes the actions of the clinicians in the cohort of sepsis and septic shock patients along with a prescribed reward function to learn optimal dosing strategies. First, we devise a reward function that in some way quantifies the effective improvement of a patient over the last hour given their state variables and the treatment actions that they received. In general, the reward function at time *t* is of the form

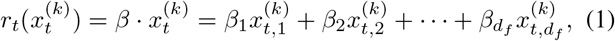

where the **x**_*t*_ is the observational data at time *t* and *β* is a vector of weights for the corresponding observations. The vector of weights *β* should be chosen with some clinical insight that is reasonable for a septic patient, i.e many of the weights could be zero, indicating that specific observational inputs are not significant in determining if a patient is improving. For this study, we compute the reward using only the following observational variables: the change in SOFA score^1^ and all of its end-organ related components (i.e., respiratory SOFA, cardiovascular SOFA, liver SOFA, and kidney SOFA), duration of hypotensive episodes, urine output over the last four hours, and a hyperbolic tangent function of MAP that assigns a positive reward to a MAP of greater than 65 mmHg. All of these variables are highly correlated with a septic patient’s status improving, making them a reasonable choice for the independent variables for the reward function. The vector of weights *β* is then treated as a hyperparameter that can be tuned using optimization techniques, which we will discuss in more detail in the section “Policy Evaluation and Causal Treatment Effects”.

#### 1) Patient State Estimation

As a first step to learning a reinforcement policy, the agent must be able to identify the “state” that the patient is in at any given point in time. We utilize a supervised approach to hidden state estimation under the assumption of Markovianity and a linear state transition model [13]. This approach obviates the need for a generative model for the observations which often include a combination of continuous and categorical variables. We assume that there are *J* possible hidden states (or *modes*) in the time series data^2^, and the likelihood function of states takes the form of a softmax classifier with parameters *α*; mapping the observations to the likelihood of the *J* latent states. The network uses a forward pass over the time series data to predict the latent states using the transition matrix *Z* and the supervised likelihood model. Learning of the model parameters is achieved by unrolling the model into a neural network and training the resulting network to find a set of states and parameters that optimizes the RL agent’s cost function (i.e., training is done end-to-end akin to a deep reinforcement learning model).

For a Hidden Markov Model, the posterior probability of the latent state at time *t* given the set of observations up to that time is given by [14]:

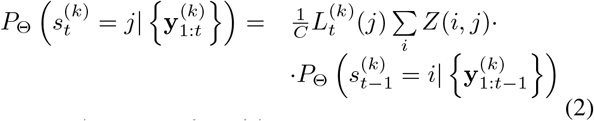

where 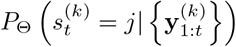 denotes the probability that the latent state *s* at time *t* is equal to *j* given the observations 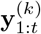, *C* is a normalizing factor, 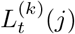 is the likelihood function that is given by 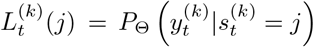 (that is, the computed probability of the observation 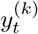 given the latent state 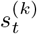 is *j*), and Θ = {*α, Z*} is the set of model parameters. In a supervised setting, the likelihood function can be replaced with a softmax classifier that will be trained along with the rest of the RL agent network, so that with each pass through the observational data, not only will the RL agent learn more effective treatments to use given the set of time series points, but the model will also be able to more accurately predict the hidden state of the patient at each time point. The network describing the estimation of the latent patient states is depicted in the lower right portion of Fig. 1. Henceforth, we call this network a switching generalized linear model (or SGLM).

**Fig. 1.**
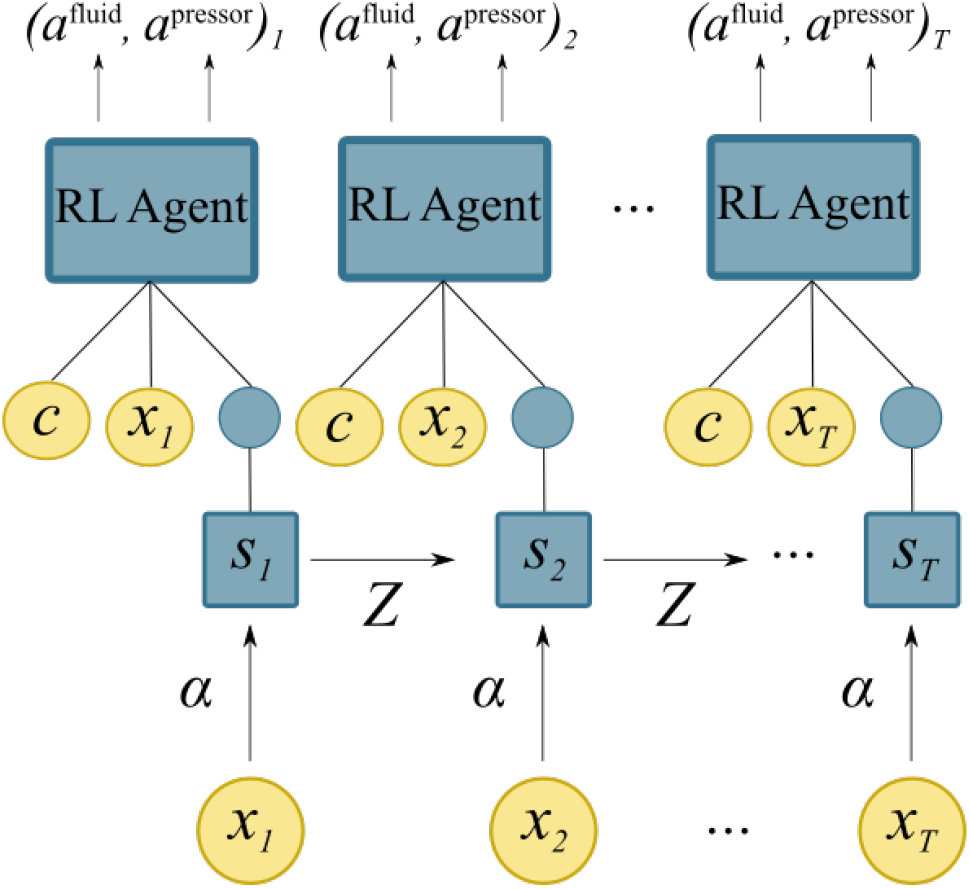
Conceptual scheme for the dosing recommendation process. The model receives the patient clinical features **x**_*t*_ from which the current state *s*_*t*_ is predicted. Then, using the predicted state, the clinical features, and covariate features **c**, the network recommends a dosing action pair (*a*^fluid^, *a*^pressor^)_*t*_. Color indicates latent states and RL policy requiring inference and learning via gradient backpropagation.

#### 2) Reinforcement Learning in Continuous State and Action Spaces

After deciphering the patient’s mode (hidden state) from the set of *J* modes, the agent can then be presented with the state, observational data, and the patient’s covariates to judge the appropriate action to be taken. Because the RL agent is not permitted to freely explore the state and action space itself, it relies on the existing patient data to learn the utility of each action. In effect, the agent treats the actions of the clinician as a “random” exploration of the state and action space. After finding an appropriate set of weights *β* that allows for computing a scalar reward from Eq. (1) the agent can be trained using the REINFORCE algorithm [15], to recommended dosing as a pair 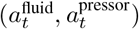 in the continuous domain. The REINFORCE network learns to maximize the expected accumulated reward of the policy under the probability distribution generated by the SGLM model,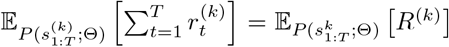. The gradient of this cost function is known to be easily approximated by

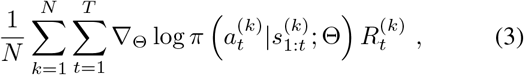

where 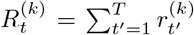 is the cumulative reward obtained following the action 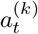, and Θ (with a slight abuse of notation) is now the combined parameters of the REINFORCE network and the SGLM state estimator. Essentially, the agent observes how state action pairs are rewarded (over an entire patient trajectory), and changes in the REINFORCE network policy are informed directly by the magnitude and sign of the reward for a given state action pair. Put more simply, the network is encouraged to take actions in a specific state that yield more positive accumulated long-term rewards, and discouraged to take actions that yield negative accumulated long-term rewards.

Figure 1 summarizes the RL agent network structure. Observations *x*_*t*_ are used to predict the state *s*_*t*_, which is then used to predict the next state and also fed into the Reinforcement Learning network (along with the observations *x*_*t*_ and covariates *c*). In practice, we observe that the RL agent performs better when having access to both *x*_*t*_ and *s*_*t*_, which is likely due to the federated information flow induced by this network architecture [16].

### C. Policy Evaluation and Causal Treatment Effects

Training a reinforcement learning agent to make sequential decisions from historic data is insufficient to make a claim that the agent is *useful* in practice. Since the data distribution is directly affected by a new policy (*π*^*a*^), and it is impossible to determine exactly *how* the agent’s action at a given time point will affect the environment downstream if it differs from the existing policy (*π*^*c*^). However, a number of approaches have been proposed for off-policy evaluations in such setting [17], [18]. These methods rely on learning a model of the system from the existing data and using importance sampling to correct the mismatch between the distributions of the system trajectory induced by the new and the existing policies. These methods correct the mismatch between the distributions by assigning no (or almost no) weight to the trajectories where the agent and real clinician recommend different actions. Additionally, these methods focus on the internal rewards used to train the new policy, whereas our goal is to assess the potential effect of a new policy on the long-term outcome of a patient (e.g., in-hospital or 30-days survival). Therefore, we propose to estimate the causal effects associated with the observed deviations from the RL policy in the clinical practice and the long-term outcomes of patients.

The propensity score was first introduced as a way to estimate the propensity of a subject (here a patient) in a study to get exposed to an intervention (here a treatment policy) based on observed covariates [19]. Propensity scores were originally presented as a way to make causal inferences about the relationship between covariates and binary treatment levels. The Generalized Propensity Score (GPS) method was developed to extend causal inference to the case of multiple and continuous treatments. More formally, the GPS is the conditional probability that an individual receives treatment *t* ∈*T* = {*τ*_1_, *τ*_2_, …, *τ*_*n*_} given covariates *x*. Although the GPS has been extended to continuous treatment spaces [20], [21] by means of estimating the conditional density function, we elect to limit our attention to the case of discrete treatment levels for the sake of simplicity.

To define the treatment levels, we first compute the distance (*d*) between the recommended policy, *π*(*a* |*s, θ*), and the policy implemented by the clinicians in the ICU, *π*_*c*_(*x*). This distance, 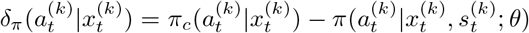 represents how different the agent’s recommended policy is for patient *k* at time *t* from the clinician’s policy. We then partition this policy difference space into *m*^2^ bins (*m* for intravenous fluids and *m* for vasopressors). This space defines the treatment levels for the purpose of causal treatment effects assessment and policy evaluation. Note that a treatment level of zero (*d*_*t*_ = 0) for intravenous fluids and vasopressors corresponds to the case where the clinical actions exactly match the RL recommended actions.

The propensity to receive each different treatment level *τ*_*i*_ (given by *GPS*(*τ*_*i*_ |*x*)) can then be treated as a softmax classifier (namely the GPS-network), where given a set of observations at a given time, the model returns the patients’ propensity to receive a given treatment level (including the need to deviate from the clinician’s dosage). This propensity score enables us to assess the effects of a given treatment level on the patient outcome (*Y*), through causal inference with discrete treatment levels and binary outcomes [20], [22]. Creating an averaged dose response function (ADRF) by mapping the received treatment levels and the associated GPS to the outcome of interest (e.g., hospital survival) allows us to explore counterfactual reasoning for vasopressor and fluid dosing. That is, how might a patient’s outcome change if the patient is given different levels of treatment than the hospital policy recommends? This is one of the key questions in the application of RL to sequential dosing of medications in many clinical settings. In this work, we utilize a neural network with a logistic classification layer (namely the ADRF-network, which can be trained as a normal logistic classifier) to model the ADRF function. Given a set of clinical observations and counterfactual levels of treatment, first the GPS-network is used to map the observations to the GPS score for the different treatment levels. Next, the GPS scores and the counterfactual treatments are fed into the ADRF-network model to estimate the expected effect size, that is, the difference between the ADRF-network output evaluated at the counterfactual (*d*_*t*_ = 0) and the actual 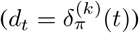 treatment levels. The corresponding cost function is defined as:

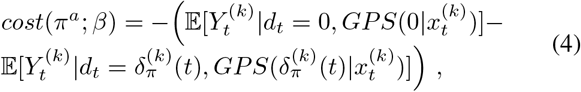

where the first term inside the parentheses on the right-hand-side of the question is the propensity-adjusted expected survival assuming the clinician’s action was exactly the same as the RL agents suggestion, and the second term is the propensity adjusted expected survival under the observed level of treatment. Note, that minimization of the cost function in Eq. (4) can be accomplished by 1) optimizing the reward function in Eq. (1), and 2) improving the learned RL policy via Eq. (3). To accomplish the former we use Bayesian Optimization [23] to optimize the reward function parameters *β*. Next, given the reward function, the REINFORCE algorithm is used to learn an optimal dosing policy. This allows our model to be trained using both short- and long-term rewards, which creates a more balanced dosing agent. A high-level algorithm for training the reward function parameters and RL Agent/Patient State Estimator is given in Algorithm 1.

#### Algorithm 1 Train Dosing Agent and Reward

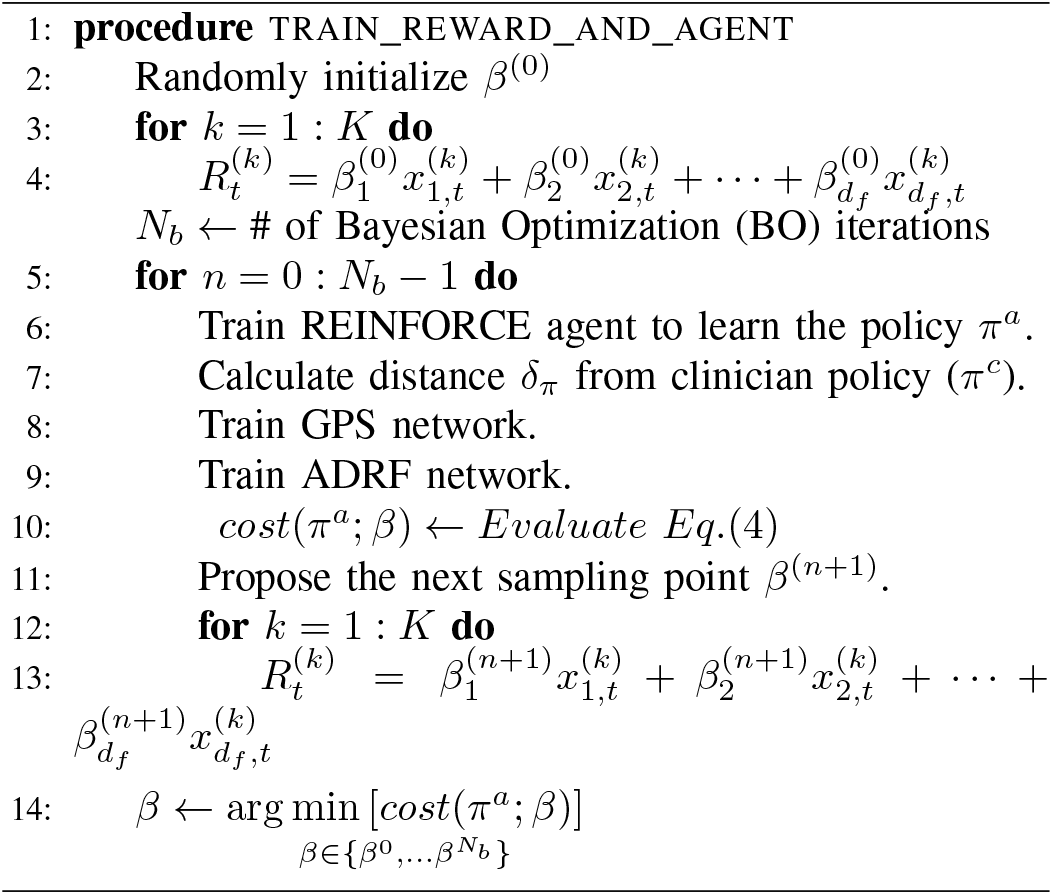

#### 1) Calibrating the Dose Response Function

Predicting the average response to a treatment can have many pitfalls that one should be cognizant of, and treat with care. The most significant of which is the potential for a classifier to be uncalibrated. That is, when the probabilities the classifier outputs do not adequately represent the corresponding proportions in the underlying population. For example, when predicting mortality, ideally the subset of a patient cohort that receives a probability of 0.1 should have a 10% prevalence of mortality. Often, with trained models, calibrating the outputs of the classifier can be done just by performing a shift on the outputs. One popular such shift is isotonic regression [24], [25], [26], which fits a non-decreasing curve to a set of data. Using isotonic regression, we can map the outputs of our ADRF network to their corresponding proportions in the dataset, without losing the ordering of the predictions.

To assess whether or not a classifier is calibrated, one can perform a Hosmer-Lemeshow goodness-of-fit test [27]. The H-L goodness-of-fit test amounts to using a Pearson’s chi-squared test on a test statistic that is constructed from percentile (typically decile) binning of the data. This test gives a reasonable idea as to whether or not the empirical values and predicted values feasibly represent the same underlying distribution.

When training our model, we use the H-L *p* value to determine whether or not the dose-response model is calibrated. Then, pending the results of the test calibrate the model, if necessary, using isotonic regression.

## III. Results

We trained and tested our Reinforcement Learning dosing agent on a subset of the sepsis and septic shock patients from the MIMIC III database. Figure 2 shows a time series comparision of the different dosing recommendations of the clinician (red) and the RL agent (blue) along-side a patient’s MAP and the estimated reward function. It is important to note that the RL agent’s decisions only take into account the patient’s time series, and not its previous actions (because it cannot directly influence the environment). This means that the RL agent is not recommending repeated doses of pressors necessarily, but rather suggesting that based on the current state of the patient at a given hour that pressors be given. Remarkably (as evidenced by this figure and numerous other instances in the test set), the agent learns the inverse relationship between Mean Arterial Pressure (MAP) and vasopressor dosing. Typically, vasopressors are given to patients whose blood pressure drops below 65 and can not be restored through intravenous fluids.

**Fig. 2.**
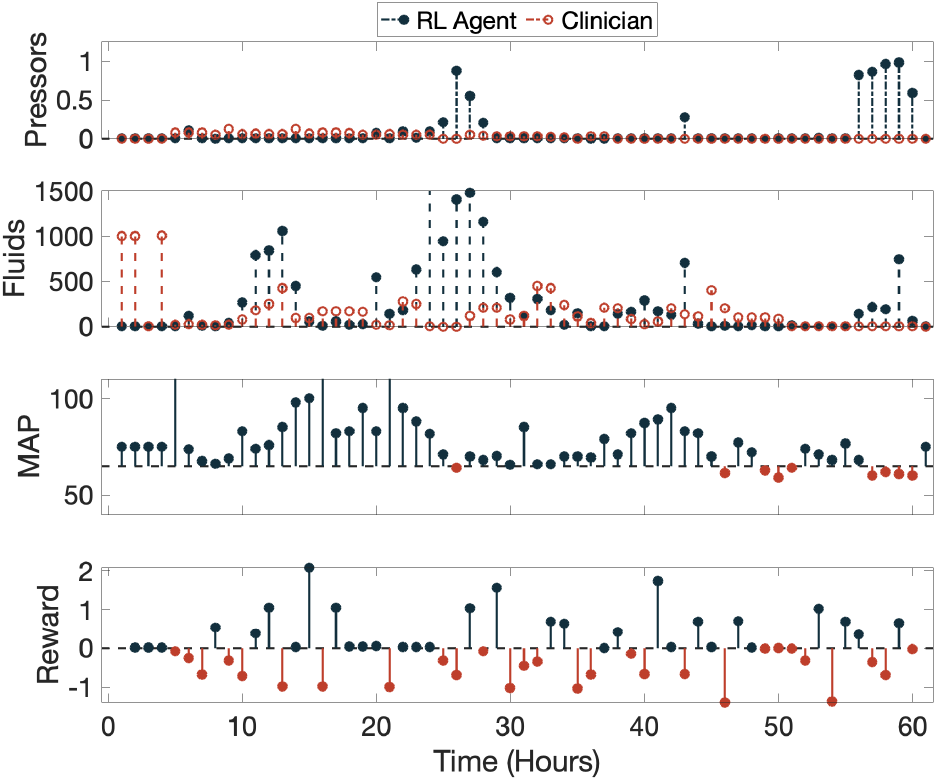
Example of pressor (top) and fluid (second from top) dosing by clinicians (red) and the RL agent (blue) alongside the patient’s MAP (second from bottom) and the calculated reward (bottom) for the clinician at each time point. In the MAP and Reward panels, red indicates MAP < 65 and reward < 0, respectively.

### A. Comparative Performance

The clinician and the RL dosing agent take noticeably different approaches to fluid resuscitation in patients. Table I outlines the key distinctions between the two policies. The RL agent tends to recommend fluids earlier and in higher amounts than the clinician, while simultaneously delaying and recommending fewer pressors. Intravenous fluids are typically the first methods clinicians use to raise blood pressure in septic patients, and it is an encouraging sign for the agent to learn this from the environment.

**TABLE I.**
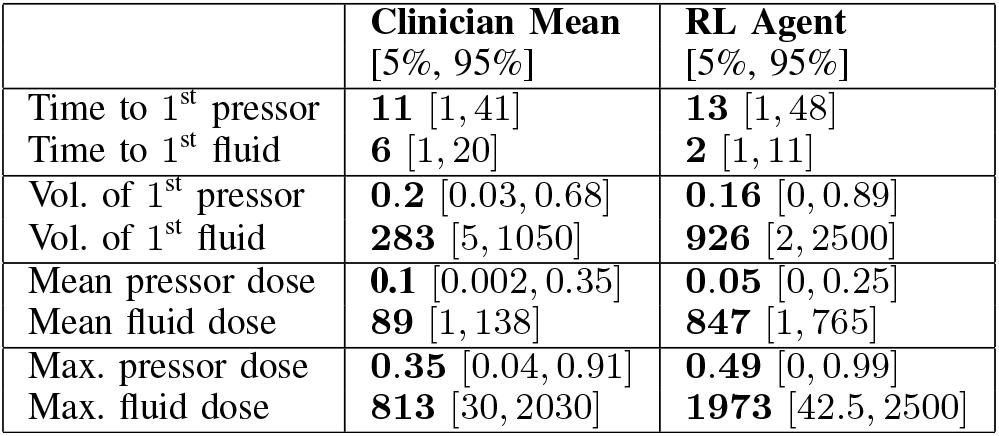
Comparison of the clinical policy and the RL Agent dosing policy.

|  | Clinician Mean<br>[5%, 95%] | RL Agent<br>[5%, 95%] |
| --- | --- | --- |
| Time to 1<sup>st</sup> pressor | 11 [1, 41] | 13 [1, 48] |
| Time to 1<sup>st</sup> fluid | 6 [1, 20] | 2 [1, 11] |
| Vol. of 1<sup>st</sup> pressor | 0.2 [0.03, 0.68] | 0.16 [0, 0.89] |
| Vol. of 1<sup>st</sup> fluid | 283 [5, 1050] | 926 [2, 2500] |
| Mean pressor dose | 0.1 [0.002, 0.35] | 0.05 [0, 0.25] |
| Mean fluid dose | 89 [1, 138] | 847 [1, 765] |
| Max. pressor dose | 0.35 [0.04, 0.91] | 0.49 [0, 0.99] |
| Max. fluid dose | 813 [30, 2030] | 1973 [42.5, 2500] |

Figure 3 shows the result of counterfactual reasoning over five different pressor and fluid distance levels. The best performance, in terms of the expected improvement in survival rate, is achieved when the clinician dosing policy closely follows the RL recommended dosing of fluids and pressors. As expected, dramatic deviations from the RL policy (represented by *δ* _*Pressor*_ and *δ* _*Fluid*_ far from 0) are not associated with a positive change in survival rate, and may even hurt the patients. Indeed, the most dramatic improvement in the survival rate occurs when the policy is close to the RL policy (indicated by *δ*_*Fluid*_ ∈ [−500, 500] and *δ*_*Pressor*_ ∈ [−0.2, 0.2]). This corresponds to a survival rate improvement of ≈3%.

**Fig. 3.**
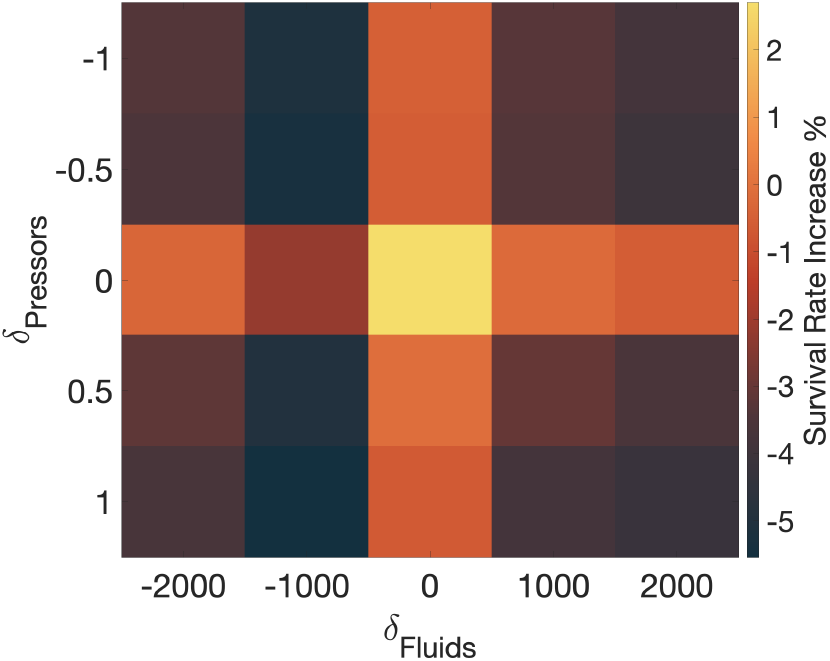
Summary of the expected improvement in survival rate for as function of deviation from the RL dosing policy (i.e., negative of the cost function in Eq. (4)). The largest survival rate increase occurs when *δ*_Fluid_ and *δ*_Pressor_ are close to zero, indicating adherence to the RL policy may result in better outcomes even after adjusting for confounding factors via proposing matching.

This improvement in performance contrasts with the performance of versions of the model i. with no reward (using Imitation Learning [28]), ii. that focus only on short-term rewards, or iii. that do not include the temporal component (by virtue of the SGLM network). The imitation learning model acts as a “mean” clinician policy, in that during training, the model sees a random sample of patient states and policies. The model that only learns from short-term rewards (for example only dosing based on a patient’s MAP value) is penalized when viewing causal treatment effects, because vasoactive drugs have potentially toxic effects [29]. Lastly, the REINFORCE model without the SGLM temporal components fails to capitalize on information from a patient’s history. Respectively, these models see a 1.3*x*% (IL), 1.2% (short-term only), and 1.9% (vanilla REINFORCE) increase in survival for the test cohort.

## IV. Discussions and Conclusion

We have shown that by combining reinforcement learning with the framework of causal treatment effects one may simultaneously learn reward functions and treatment policies that are directly tuned to optimize a long-term outcome. This enables the end-to-end training of a model that increases survival rates in the testing cohort to ≈ 3%. While the application of reinforcement learning to medication dosing in critically ill patients was previously suggested by [6], [30], and [31] in the context of sepsis and septic shock, our work presents several novel contributions.

Compared to the model proposed by Raghu, et al., our agent learns to provide treatment in a continuous action space that directly corresponds to the clinical action space, whereas Raghu, et. al. discretize their 2D action space to utilize Q-learning, which can result in a loss of treatment precision. Secondly, while Raghu, et. al. train their Q-Network and auto-encoder separately, we use end-to-end training of the state estimator and the reinforcement learning agent (REINFORCE). End-to-end training is known to provide more optimal models, as only a single cost function is optimized. Finally, Raghu, et. al. limit their reward to in-hospital mortality. Without adjusting for confounding factors, defining the reward in this way tends to bias the learned policy towards treatment strategies that clinicians use on less sick patients who have a higher chance of survival.

In another recent work [31], Komorowski, et. al. present a reinforcement learning agent trained using policy iteration [4]. This method requires the authors to discretize both the patient clinical course and covariate factors, as well as the action space. This reduction of dimensionality can reduce the richness of the information provided to a model, and subsequently affect the ability of the model to make decisions. Furthermore, Komorowski, et. al. do not limit their cohort to septic patients requiring vasopressor therapy. This skews both the state representation of their patients, as well as the dosing recommendation of their agent – their agent is much more likely to recommend minimal doses. Most significantly, the method of policy evaluation they use, importance sampling [32], biases the evaluation to ignore patient trajectories in which the AI Clinician’s policy differs from the real clinician. This ultimately predisposes their clinician to inaction in patients with a higher mortality rate. We discuss the limitations of their agent in more detail in [33]. Our methods are in stark contrast to this, as our agent is trained in a continuous state space, and our evaluation methods adjust for confounding factors using causal treatment effects.

In [32] and [9], the authors present the two primary confounds that underlie utilizing and evaluating reinforcement learning methods in a critical care setting: proper patient state representation and proper statistical evaluation of policy to compensate for confounding factors. They recommend utilizing causal inference-based methods for evaluating the policy and limiting policies that are similar to clinician policies. This second recommendation is critical in medicine. It is particularly challenging to evaluate treatment policies that are dramatically different than the clinical policy, because there are inevitably additional confounding factors that influence clinicians’ decisions when providing treatment. We have taken these recommendations and have carefully devised the first reinforcement learning algorithm in a critical care setting that properly adjusts for confounding factors, with the intention of making meaningful and informed decisions.

Our method constructs a clinically-significant reward function and then simultaneously learns to assign a state to a patient from their clinical information and recommend a treatment based on their state and covariate factors. We then estimate patients’ propensity to receive treatment and their expected response to that treatment to gauge their expected change in survival under the new treatment policy. We find that adherence to the individualized fluid and vasopressor dosing policy is significantly associated with mortality reduction of ≈ 3% in patients suffering from sepsis or septic shock, even after adjusting for severity of illness.

Our method is not without its limitations. Currently, the agent cannot be placed in a real patient treatment environment. It must learn its policy from observing clinicians exploring the action space and being rewarded by patient improvement over short and long time scales. Indeed, off-policy evaluation can be particularly challenging. In our method, we evaluate our policy using standard counterfactual reasoning in a retrospective setting, there is room to use other evaluation metrics and techniques. Additionally, we recognize that each class of vasopressors and (to a lesser degree) fluids induce a unique physiologic response. In this study we normalize both types of treatments to simplify our agent’s action space. In reality, every type of vasopressor or fluid does not carry the same utility depending on patient status and history, and are even dosed differently. The choice of clinical features to use in the reward function was based on known factors in judging a patient’s improvement when suffering from sepsis. The reward function should be analyzed more methodically to understand the roles of different clinical factors in assessing improvement in patient state; methods like functional ANOVA [34] that determine the dependence of a function on the input variables can improve the reward function and potentially add new clinically significant meaning to it.

Despite these simplifications which will be explored in future work, this precursory reinforcement learning approach marks an exciting first step towards providing real-time, individualized computer-assisted treatment of the hypotensive episodes that can accompany sepsis and septic shock.

## Data Availability

We have used the publicly available MIMIC-III (Medical information for intensive care) dataset of critically ill patients, available at https://mimic.physionet.org/

## Acknowledgments

This work was supported by the National Institutes of Health (NIH) early career development award in biomedical big data science (1K01ES025445-01A1) and NIH grant 2R01GM104987-09. The content of this article is solely the responsibility of the authors and does not necessarily represent the official views of the NIH.

## Appendix - Data Description and Extraction

This appendix contains specific details about the clinical variables used from the MIMIC III database [11] for the cohort of sepsis and septic shock patients. Vital signs and clinical variables were binned on an hourly basis. The following time-varying variables were included: heart rate, mean blood pressure, systolic blood pressure, respiratory rate, temperature, Glasgow Coma Scale (GCS), SpO2, FiO2, PaO2/FiO2 ratio, paO2, paCO2, arterial base excess, arterial pH, arterial lactate, bicarbonate (HCO3), potassium, sodium, chloride, glucose, blood urea nitrogen (BUN), creatinine, magnesium, calcium, total bilirubin, albumin, hemoglobin, white blood cell (WBC) count, platelets count, urine output, and Sequential Organ Failure Assessment score (SOFA). Additionally, we include the total fluid input administered and median vasopressor dosage in a given hour. We used carry-forward to fill in missing values on an hourly basis, except for input and output events (i.e. urine output and amount of fluids administered) and vasopressor dosage. At each time step, the mean blood pressure, fluids and vasopressor dosage from the previous three time-steps were also included to infer patient states.

The following static variables were included: gender, age, Elixhauser co-morbidity index, and five severity-of-illness scores computed using data from the first 24-hours patients were in the ICU, including SAPS-II, APS-III, OASIS, SOFA, and Charlson Comorbidity Index (CCI).

There are many different types of vasopressors that are dosed differently. Vasopressors are standardized using the relative strength to norepinephrine (aka noradrenaline or norad). The vasopressors included: norad (or norepinephrine or levophed), epi (adrenaline), vasopressin, phenyl, and dopa. The unit of measurement is mcg/kg/minute except for vasopressin (rate is expressed as units/minute). The standardization step involves multiplying the rate of each vasopressor by a scaling constant based on usual dosing of each drug. Norad is typically dosed at 0-1 mcg/kg/min. If multiple vasopresors were used in the same time bin, the median dose is reported.

## Footnotes

1 Sequential Organ Failure Assessment (SOFA) scores are often used by clinicians to quickly assess organ-specific deterioration [12].

2 In this study, we assumed *J* is a hyper parameter for Bayesian Optimization

## References

[1] M. Singer, C. S. Deutschman, C. W. Seymour, M. Shankar-Hari, D. Annane, M. Bauer, R. Bellomo, G. R. Bernard, J.-D. Chiche, C. M. Coopersmith, R. S. Hotchkiss, M. M. Levy, J. C. Marshall, G. S. Martin, S. M. Opal, G. D. Rubenfeld, T. van der Poll, J.-L. Vincent, and D. C. Angus, “The Third International Consensus Definitions for Sepsis and Septic Shock (Sepsis-3),” JAMA, vol. 315, no. 8, pp. 801–810, Feb. 2016. [Online]. Available: https://www.ncbi.nlm.nih.gov/pmc/articles/PMC4968574/

[2] C. Rhee, R. Dantes, L. Epstein, D. J. Murphy, C. W. Seymour, T. J. Iwashyna, S. S. Kadri, D. C. Angus, R. L. Danner, A. E. Fiore, J. A. Jernigan, G. S. Martin, E. Septimus, D. K. Warren, A. Karcz, C. Chan, J. T. Menchaca, R. Wang, S. Gruber, and M. Klompas, “Incidence and Trends of Sepsis in US Hospitals Using Clinical vs Claims Data, 2009-2014,” JAMA, vol. 318, no. 13, pp. 1241–1249, Oct. 2017. [Online]. Available: http://jamanetwork.com/journals/jama/fullarticle/REFj

[3] M. Shankar-Hari, G. S. Phillips, M. L. Levy, C. W. Seymour, X. Liu, C. S. Deutschman, D. C. Angus, G. D. Rubenfeld, and M. Singer, “Developing a New Definition and Assessing New Clinical Criteria for Septic Shock: For the Third International Consensus Definitions for Sepsis and Septic Shock (Sepsis-3),” JAMA, vol. 315, no. 8, pp. 775–787, Feb. y2016. x[Online]. Available: http://jamanetwork.com/journals/jama/fullarticle/2492876

[4] R. S. Sutton, A. G. Barto et al., Reinforcement learning: An introduction. MIT press, 1998.

[5] B. L. Moore, L. D. Pyeatt, V. Kulkarni, P. Panousis, K. Padrez, and A. G. Doufas, “Reinforcement learning for closed-loop propofol anesthesia: a study in human volunteers,” The Journal of Machine Learning Research, vol. 15, no. 1, pp. 655–696, 2014.

[6] S. Nemati, M. M. Ghassemi, and G. D. Clifford, “Optimal medication dosing from suboptimal clinical examples: A deep reinforcement learning approach,” in Engineering in Medicine and Biology Society (EMBC), 2016 IEEE 38th Annual International Conference of the. IEEE, 2016, pp. 2978–2981.

[7] M. K. Bothe, L. Dickens, K. Reichel, A. Tellmann, B. Ellger, M. Westphal, and A. A. Faisal, “The use of reinforcement learning algorithms to meet the challenges of an artificial pancreas,” Expert review of medical devices, vol. 10, no. 5, pp. 661–673, 2013.

[8] N. Prasad, L.-F. Cheng, C. Chivers, M. Draugelis, and B. E. Engelhardt, “A reinforcement learning approach to weaning of mechanical ventilation in intensive care units,” arXiv preprint arXiv:1704.06300, 2017.

[9] O. Gottesman, F. Johansson, M. Komorowski, A. Faisal, D. Sontag, F. Doshi-Velez, and L. A. Celi, “Guidelines for reinforcement learning in healthcare,” Nature medicine, vol. 25, no. 1, pp. 16–18, 2019.

[10] V. Mnih, K. Kavukcuoglu, D. Silver, A. A. Rusu, J. Veness, M. G. Bellemare, A. Graves, M. Riedmiller, A. K. Fidjeland, G. Ostrovski et al., “Human-level control through deep reinforcement learning,” Nature, vol. 518, no. 7540, p. 529, 2015.

[11] A. E. Johnson, T. J. Pollard, L. Shen, L. Lehman, M. Feng, M. Ghassemi, B. Moody, P. Szolovits, L. Celi, and M. R.G., “MIMIC-III, a freely accessible critical care database,” Scientific Data, vol. 3, no. 160035, 2016.

[12] J. L. Vincent, A. de Mendoncça, F. Cantraine, R. Moreno, J. Takala, P. M. Suter, C. L. Sprung, F. Colardyn, and S. Blecher, “Use of the SOFA score to assess the incidence of organ dysfunction/failure in intensive care units: results of a multicenter, prospective study. Working group on “sepsis-related problems” of the European Society of Intensive Care Medicine,” Critical Care Medicine, vol. 26, no. 11, pp. 1793–1800, Nov. 1998.

[13] S. Nemati and R. Adams, “Identifying outcome-discriminative dynamics in multivariate physiological cohort time series,” Advanced State Space Methods for Neural and Clinical Data, vol. 283, 2015.

[14] K. P. Murphy, Machine Learning: A Probabilistic Perspective. The MIT Press, 2012.

[15] R. J. Williams, “Simple statistical gradient-following algorithms for connectionist reinforcement learning,” Machine learning, vol. 8, no. 3-4, pp. 229–256, 1992.

[16] R. K. Srivastava, K. Greff, and J. Schmidhuber, “Highway networks,” arXiv preprint arXiv:1505.00387, 2015.

[17] N. Jiang and L. Li, “Doubly robust off-policy value evaluation for reinforcement learning,” arXiv preprint arXiv:1511.03722, 2015.

[18] M. Farajtabar, Y. Chow, and M. Ghavamzadeh, “More robust doubly robust off-policy evaluation,” arXiv preprint arXiv:1802.03493, 2018.

[19] P. R. Rosenbaum and D. B. Rubin, “The central role of the propensity score in observational studies for causal effects,” Biometrika, vol. 70, no. 1, pp. 41–55, 1983.

[20] K. Hirano and G. W. Imbens, “The propensity score with continuous treatments,” Applied Bayesian modeling and causal inference from incomplete-data perspectives, vol. 226164, pp. 73–84, 2004.

[21] P. C. Austin, “Assessing the performance of the generalized propensity score for estimating the effect of quantitative or continuous exposures on binary outcomes,” Statistics in medicine, vol. 37, no. 11, pp. 1874–1894, 2018.

[22] K. Imai and D. A. Van Dyk, “Causal inference with general treatment regimes: Generalizing the propensity score,” Journal of the American Statistical Association, vol. 99, no. 467, pp. 854–866, 2004.

[23] J. Snoek, H. Larochelle, and R. P. Adams, “Practical bayesian optimization of machine learning algorithms,” in Advances in neural information processing systems, 2012, pp. 2951–2959.

[24] N. Chakravarti, “Isotonic median regression: a linear programming approach,” Mathematics of operations research, vol. 14, no. 2, pp. 303–308, 1989.

[25] P. Mair, K. Hornik, and J. de Leeuw, “Isotone optimization in r: pool-adjacent-violators algorithm (pava) and active set methods,” Journal of statistical software, vol. 32, no. 5, pp. 1–24, 2009.

[26] J. De Leeuw, “Correctness of kruskal’s algorithms for monotone regression with ties,” Psychometrika, vol. 42, no. 1, pp. 141–144, 1977.

[27] D. W. Hosmer Jr, S. Lemeshow, and R. X. Sturdivant, Applied logistic regression. John Wiley & Sons, 2013, vol. 398.

[28] S. Ross, G. Gordon, and D. Bagnell, “A reduction of imitation learning and structured prediction to no-regret online learning,” in Proceedings of the fourteenth international conference on artificial intelligence and statistics, 2011, pp. 627–635.

[29] M. N. Bangash, M.-L. Kong, and R. M. Pearse, “Use of inotropes and vasopressor agents in critically ill patients,” British journal of pharmacology, vol. 165, no. 7, pp. 2015–2033, 2012.

[30] A. Raghu, M. Komorowski, L. A. Celi, P. Szolovits, and M. Ghassemi, “Continuous state-space models for optimal sepsis treatment-a deep reinforcement learning approach,” arXiv preprint arXiv:1705.08422, 2017.

[31] M. Komorowski, L. A. Celi, O. Badawi, A. C. Gordon, and A. A. Faisal, “The artificial intelligence clinician learns optimal treatment strategies for sepsis in intensive care,” Nature Medicine, vol. 24, no. 11, p. 1716, 2018.

[32] O. Gottesman, F. Johansson, J. Meier, J. Dent, D. Lee, S. Srinivasan, L. Zhang, Y. Ding, D. Wihl, X. Peng et al., “Evaluating reinforcement learning algorithms in observational health settings,” arXiv preprint arXiv:1805.12298, 2018.

[33] R. Jeter, C. Josef, S. Shashikumar, and S. Nemati, “Does the “artificial intelligence clinician” learn optimal treatment strategies for sepsis in intensive care?” arXiv preprint arXiv:1902.03271 [cs.AI], 2019.

[34] F. Hutter, H. Hoos, and K. Leyton-Brown, “An efficient approach for assessing hyperparameter importance,” in Proceedings of International Conference on Machine Learning 2014 (ICML 2014), Jun. 2014, p. 754–762.

